# Joint engagement is associated with greater development of language and sensory awareness in children with autism

**DOI:** 10.1101/2022.05.25.22275584

**Authors:** Andrey Vyshedskiy, Edward Khokhlovich

## Abstract

The effect of joint engagement in 2- to 6-year-old children with ASD was investigated in the largest and the longest observational study to-date. Parents assessed the development of 12081 children quarterly for three years on five subscales: receptive language, expressive language, sociability, sensory awareness, and health. Longer duration of time spent with an adult actively involved in the same activity was associated with improved trajectory of receptive language, expressive language, and sensory awareness. On the annualized basis, the high-joint-engagement group (3 hours or more of joint engagement per day) improved their combinatorial receptive language 1.4-times faster (*p*=0.0019), expressive language 1.5-times faster (p<0.0001), and sensory awareness 1.5-times faster (*p*=0.0248) than the low-joint-engagement group (1 hour or less joint engagement per day). The difference in the sociability and the health scores at the end of 3-year study was insignificant. This study confirms the importance of ASD children spending more time with adults actively involved in the same activity and highlights the need to include joint engagement as a target for intervention with this population.

**Lay summary:** Parents of 2- to 6-year-old children with ASD assessed the development of 12081 children quarterly for three years. Longer duration of time spent with an adult actively involved in the same activity was associated with improved developmental trajectory. This study confirms the importance of ASD children spending more time with adults actively involved in the same activity and highlights the need to include joint engagement as a target for intervention with this population.

## Introduction

Time spent with an adult actively involved in the same activity is known as joint engagement (Adamson et al., 2004). While children on the spectrum often have no trouble entertaining themselves, such solitary activities do not favor language acquisition which depends on social interactions and complex dialogic conversations (Mayberry, 2002; Romeo et al., 2018; Vyshedskiy et al., 2017). Common social interactions include parent and child playing together with toys or reading a book – activities when they have opportunities to communicate both verbally and nonverbally. Increasing joint engagement between parent and child facilitates children’s language learning and is the target of many ASD early intervention programs (Kasari et al., 2010; Shih et al., 2021). Most studied early interventions targeting joint engagement include *Joint Attention, Symbolic Play, Engagement, and Regulation* (JASPER) (Shire et al., 2017), *Early Start Denver Model* (ESDM)(Dawson et al., 2010), and *Preschool Autism Communication Trial* (PACT)(Pickles et al., 2016).

The strategies in JASPER promote joint attention skills, such as looking between people and objects, showing, and pointing to show; modeling symbolic play; and regulation of self-stimulatory behaviors that interfere with learning. Over 10 clinical trials investigated the effect of JASPER. In the largest JASPER study Shire et al. randomly assigned 113 two- to three-year-old children with a diagnosis of ASD to either a JASPER treatment group or a treatment-as-usual (TAU) group (Shire et al., 2017). The JASPER treatment group received 10-weeks of 30 min every day of individual support from teaching assistants which focused on engaging children by creating play routines through imitation of and modeling new play acts. The wait-listed TAU group received a program to improve social skills through music and movement activities. Significant time by treatment interactions were found (with effect size Cohen’s f): children in the JASPER group spent more time in child initiated joint engagement than TAU children (F(1,70) = 46.13, p < .001, ES = 0.81). Children in the JASPER group also made significant gains in joint attention and play skills and maintained these improvements at 1 month follow-up.

The ESDM intervention guides parents and therapists to use play to build positive and fun relationships with children. Through play and joint activities, the child is encouraged to boost language, social and cognitive skills. Based on a meta analysis of 12 ESDM studies administered to the total of 640 children with ASD, Fuller et al. reported the aggregated effect size of 0.357 (p = 0.024; calculated using a robust variance estimation meta-analysis) (Fuller et al., 2020). This effect size was moderate with a statistically significant overall weighted average that favored ESDM recipients. The biggest improvements were in cognition (g = 0.412) and language (g = 0.408). No statistically significant effects were observed in autism severity, adaptive behavior, sociability, and repetitive behaviors.

The PACT intervention aims to increase parental responsiveness to child communication using video feedback. The longest-running PACT study was reported by Green et al. (Green et al., 2010). The researchers randomly assigned 152 ASD children, two- to five-years-of-age, to PACT or TAU. At the six year follow-up (participants mean age 10.5±0·8 years) the PACT intervention group showed a better Autism Diagnostic Observation Schedule (ADOS) comparative severity score (CSS) of 7.3±2.0 compared to the TAU group score of 7.8±1.8 (Pickles et al., 2016). The difference in the Language composite score of six subscales was 84.8±38.6 in the PACT group compared to 80.0±40.0 in the TAU group. The difference in conversational turns in dyadic interactions was 28.3±24.4 in the PACT group compared to 26.2±19.4 in the TAU group.

While the importance of joint engagement for children’s development is clear, we sought to contribute data from a longitudinal epidemiological approach involving thousands of participants. In this manuscript we report the effect of joint engagement in a group of ASD children who used a free language training app that invited parents to complete their child’s evaluation every three months (Dunn, Elgart, Lokshina, Faisman, Khokhlovich, et al., 2017b, 2017a; Dunn, Elgart, Lokshina, Faisman, Waslick, et al., 2017; Vyshedskiy et al., 2020; Vyshedskiy & Dunn, 2015). Parents assessed the development of 12081 ASD children, two- to six-years-of-age, quarterly for three years on five subscales: receptive language, expressive language, sociability, sensory awareness, and health. To assess the effect of joint engagement, we compared participants who reported 3 hours or more of joint engagement per day to participants who reported on average 1 or less hours of joint engagement per day. Longer duration of joint engagement was associated with improved trajectory of combinatorial receptive language, expressive language, and sensory awareness.

## Methods

### Participants

Participants were users of a language therapy app that was made available gratis at all major app stores in September 2015. Once the app was downloaded, caregivers were asked to register and to provide demographic details, including the child’s diagnosis and age. Caregivers consented to anonymized data analysis and completed the Autism Treatment Evaluation Checklist (ATEC) (Rimland & Edelson, 1999), an evaluation of the receptive language using the Mental Synthesis Evaluation Checklist (MSEC) (Braverman et al., 2018), as well as the Screen Time assessment and the Diet and Supplements assessment. The first evaluation was administered approximately one month after the download. The subsequent evaluations were administered at approximately three-month intervals. To enforce regular evaluations, the app became unusable at the end of each three-month interval and parents were required to complete an evaluation to regain its functionality.

#### Inclusion criteria

Inclusion criteria were identical to our previous studies of this population (Fridberg et al., 2021; Mahapatra, Khokhlovich, et al., 2018; Vyshedskiy et al., 2020). Specifically, we selected participants based on the following criteria:

1. Consistency: Participants must have filled out at least three ATEC evaluations and the interval between the first and the last evaluation was six months or longer.
2. Diagnosis: ASD diagnosis at the end of the study. Children without ASD diagnosis at the end of the study were excluded. Other diagnostic options included: Suspected ASD, Mild Language Delay, Pervasive Developmental Disorder, Attention Deficit Disorder, Social Communication Disorder, Specific Language Impairment, Apraxia, Sensory Processing Disorder, Down Syndrome, Lost Diagnosis of ASD or PDD, Other Genetic Disorder, Normally Developing Child. Most app users (82%) reported ASD diagnosis by their last evaluation. A good reliability of such parent-reported diagnosis was previously demonstrated (Jagadeesan et al., 2022). Furthermore, the parent-reported ASD diagnosis was supported by absolute values of ATEC scores. Average initial ATEC total score was 69.0 ± 26.2, which corresponds to severe ASD as delineated earlier (Mahapatra, Vyshedsky, et al., 2018) and Table 1.

#### Exclusion criteria

1. Maximum age: Participants older than six years of age at the time of their first evaluation were excluded from this study.
2. Minimum age: Participants who completed their first evaluation before the age of two years were excluded from this study.

After excluding participants that did not meet these criteria, there were 12081 total participants.

**Table 1.**
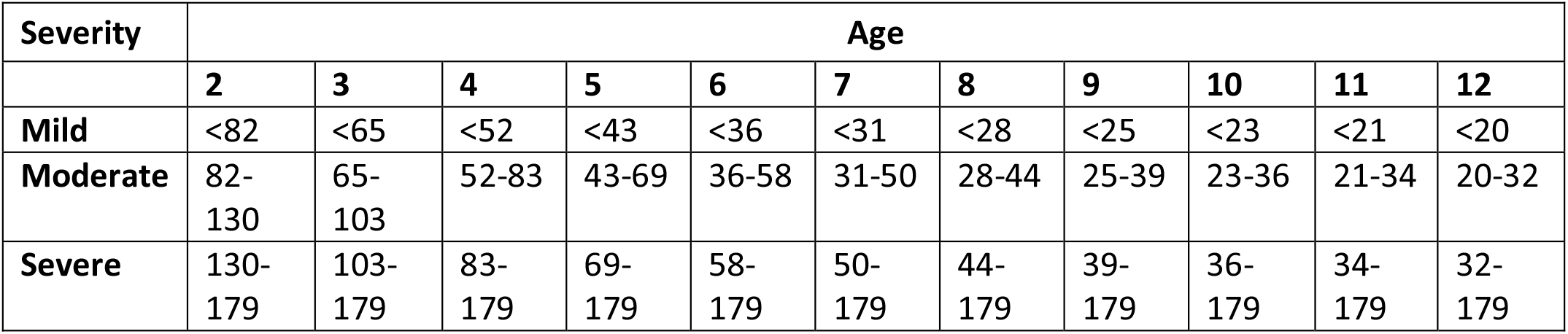
**Approximate relationship between ATEC total score, age, and ASD severity as described in Mahapatra *et al*.** (Mahapatra, Khokhlovich, et al., 2018). **At any age, a greater ATEC score indicates greater ASD severity.**

| Severity | Age |  |  |  |  |  |  |  |  |  |  |
| --- | --- | --- | --- | --- | --- | --- | --- | --- | --- | --- | --- |
|  | 2 | 3 | 4 | 5 | 6 | 7 | 8 | 9 | 10 | 11 | 12 |
| **Mild** | <82 | <65 | <52 | <43 | <36 | <31 | <28 | <25 | <23 | <21 | <20 |
| **Moderate** | 82-130 | 65-103 | 52-83 | 43-69 | 36-58 | 31-50 | 28-44 | 25-39 | 23-36 | 21-34 | 20-32 |
| **Severe** | 130-179 | 103-179 | 83-179 | 69-179 | 58-179 | 50-179 | 44-179 | 39-179 | 36-179 | 34-179 | 32-179 |

### Time with an adult actively involved in the same activity

Participants were required to respond to the question: “How much time, on average, does your child spend with an adult actively involved in the same activity (talking, reading, playing, therapy) each day”? To assess the effect of joint engagement, we compared participants who indicated that they spend on average 1 hour or less per day (N=2272; the low joint engagement group, abbreviated lowJE) to participants who reported on average 3 or more hours of joint engagement (N=2578; the high joint engagement group, abbreviated highJE).

### Evaluations

A caregiver-completed Autism Treatment Evaluation Checklist (ATEC) (Rimland & Edelson, 1999) and Mental Synthesis Evaluation Checklist (MSEC) (Braverman et al., 2018) were used to track child development. The ATEC questionnaire is comprised of four subscales: 1) Speech/Language/Communication, 2) Sociability, 3) Sensory/Sensory awareness, and 4) Physical/Health/Behavior. The first subscale, Speech/Language/Communication, contains 14 items and its score ranges from 0 to 28 points. The Sociability subscale contains 20 items within a score range of 0 to 40 points. The third subscale, referred here as the Sensory awareness subscale, has 18 items and score range from 0 to 36 points. The fourth subscale, referred here as the Health subscale, contains 25 items and score range from 0 to 75 points. The scores from each subscale are combined in order to calculate a Total Score, which ranges from 0 to 179 points. A lower score indicates lower severity of ASD symptoms and a higher score indicates more severe symptoms of ASD. ATEC is not a diagnostic checklist and it was designed to evaluate the effectiveness of treatment (Rimland & Edelson, 1999). Therefore, ASD severity can only have an approximate relationship with the total ATEC score and age. Table 1 lists approximate ATEC total score as related to ASD severity and age as described in Mahapatra *et al*. (Mahapatra, Khokhlovich, et al., 2018).

ATEC was selected as a tool since it is one of the few measures validated to evaluate treatment effectiveness. In contrast, another popular ASD assessment tool, Autism Diagnostic Observation Schedule or ADOS, (Lord et al., 2000) has only been validated as a diagnostic tool. Various studies confirmed the validity and reliability of ATEC (Geier et al., 2013; Jarusiewicz, 2002) and several trials confirmed ATEC’s ability to longitudinally measure changes in participant performance (Charman et al., 2004; Klaveness et al., 2013; Magiati et al., 2011; Mahapatra, Khokhlovich, et al., 2018). Moreover, ATEC has been used as a primary outcome measure for a randomized controlled trial of iPad-based intervention for ASD, named “Therapy Outcomes By You” or TOBY, and it was noted that ATEC possesses an “internal consistency and adequate predictive validity” (Whitehouse et al., 2017). These studies support the effectiveness of ATEC as a tool for longitudinal tracking of symptoms and assessing changes in ASD severity.

### Expressive language assessment

The ATEC Speech/Language/Communication subscale includes the following questions: 1) Knows own name, 2) Responds to ‘No’ or ‘Stop’, 3) Can follow some commands, 4) Can use one word at a time (No!, Eat, Water, etc.), 5) Can use 2 words at a time (Don’t want, Go home), 6) Can use 3 words at a time (Want more milk), 7) Knows 10 or more words, 8) Can use sentences with 4 or more words, 9) Explains what he/she wants, 10) Asks meaningful questions, 11) Speech tends to be meaningful/relevant, 12) Often uses several successive sentences, 13) Carries on fairly good conversation, and 14) Has normal ability to communicate for his/her age. With the exception of the first three items, all items in the ATEC subscale 1 primarily measure expressive language. Accordingly, the ATEC subscale 1 is herein referred to as the Expressive Language subscale to distinguish it from the Receptive Language subscale tested by the MSEC evaluation.

### Receptive language assessment

The MSEC evaluation was designed to be complementary to ATEC in measuring complex receptive language. Out of 20 MSEC items, those that directly assess receptive language are the following: 1) Understands simple stories that are read aloud; 2) Understands elaborate fairy tales that are read aloud (i.e., stories describing FANTASY creatures); 3) Understands some simple modifiers (i.e., green apple vs. red apple or big apple vs. small apple); 4) Understands several modifiers in a sentence (i.e., small green apple); 5) Understands size (can select the largest/smallest object out of a collection of objects); 6) Understands possessive pronouns (i.e. your apple vs. her apple); 7) Understands spatial prepositions (i.e., put the apple ON TOP of the box vs. INSIDE the box vs. BEHIND the box); 8) Understands verb tenses (i.e., I will eat an apple vs. I ate an apple); 9) Understands the change in meaning when the order of words is changed (i.e., understands the difference between ‘a cat ate a mouse’ vs. ‘a mouse ate a cat’); 10) Understands explanations about people, objects or situations beyond the immediate surroundings (e.g., “Mom is walking the dog,” “The snow has turned to water”); MSEC consists of 20 questions within a score range of 0 to 40 points; similarly to ATEC, a lower MSEC score indicates a better developed receptive language.

The psychometric quality of MSEC was tested with 3,715 parents of ASD children (Braverman et al., 2018). Internal reliability of MSEC was good (Cronbach’s alpha > 0.9). MSEC exhibited adequate test– retest reliability, good construct validity, and good known group validity as reflected by the difference in MSEC scores for children of different ASD severity levels. MSEC norms have been reported earlier (Arnold & Vyshedskiy, 2022).

To simplify interpretation of figure labels, the subscale 1 of the ATEC evaluation is herein referred to as the Expressive Language subscale and the MSEC scale is referred to as the Receptive Language subscale.

### Statistical analysis

The framework for the evaluation of score changes over time has been earlier explained in minute detail (Mahapatra, Khokhlovich, et al., 2018; Vyshedskiy et al., 2020). In short, the concept of a “Visit” was developed by dividing the three-year-long observation interval into 3-month periods. All evaluations were mapped into 3-month-long bins with the first evaluation placed in the first bin. When more than one evaluation was completed within a bin, their results were averaged to calculate a single number representing this 3-month interval. Thus, we had 12 quarterly evaluations for both highJE and lowJE groups.

It was then hypothesized that there was a two-way interaction between pretend-play-group and Visit. Statistically, this hypothesis was modeled by applying the Linear Mixed Effect Model with Repeated Measures (MMRM), where a two-way interaction term was introduced to test the hypothesis. The model (Endpoint ∼ Baseline + Gender + Severity + Sleep-Problem-Group * Visit) was fit using the R Bioconductor library of statistical packages, specifically the “nlme” package. The subscale score at baseline, as well as gender and severity were used as covariates. Conceptually, the model fits a plane into n-dimensional space. This plane considers a complex variability structure across multiple visits, including baseline differences. Once such a plane is fit, the model calculates Least Squares Means (LS Means) for each subscale and group at each visit. The model also calculates LS Mean differences between the groups at each visit.

In preparation for statistical analysis, participants in the lowJE group were matched to those in the highJE group using propensity score (Schneider et al., 2007) based on age, gender, expressive language, receptive language, sociability, sensory awareness, and health at the 1^st^ evaluation (baseline). The number of matched participants was 2272 in each group, Table 2.

**Table 2.** **Demographic data of participant pool.**

|  | **High Joint Engagement** | **Low Joint Engagement** |
| --- | --- | --- |
| **Number of participants** | 2508 | 2272 |
| **Number of matched participants** | 2272 | 2272 |
| **Age at baseline** | 3.6±1.0 | 3.8±1.0 |
| **Male Gender** | 78% | 76% |

## Results

To assess joint engagement, participants were required to respond to the question: “How much time, on average, does your child spend with an adult actively involved in the same activity (talking, reading, playing, therapy) each day?” High-joint-engagement participants (highJE, N=2578) who reported on average 3 or more hours of joint engagement per day were compared to low-joint-engagement participants who indicated that they spend on average 1 hour or less per day (lowJE, N=2272). In preparation for statistical analysis, children in the lowJE group were matched to those in the highJE group using propensity score (Schneider et al., 2007) based on age, gender, expressive language, receptive language, sociability, sensory awareness, and health at the 1^st^ evaluation (baseline). The number of matched participants was 2272 out of 2272 in the lowJE group and 2272 out of 2578 in the highJE group, Table 2.

We then analyzed trajectories of children development on five subscales: Receptive Language, Expressive Language, Sociability, Sensory Awareness, and Health. On the Receptive Language subscale, the average improvement in the highJE group over 36 months was 9.08 points (SE=0.49, p<0.0001) compared to 6.38 points (SE=0.51, p<0.0001) in the lowJE group, Figure 1A, Table 3, Table S1. The difference in the highJE group relative to the lowJE group at Month 36 was statistically significant: -2.17 points (SE=0.7, p=0.0019). The negative difference (marked in the Table 3 as “highJE - lowJE”) indicates that the lowJE group had greater scores at Month 36 and, therefore, more severe symptoms.

**Table 3.**
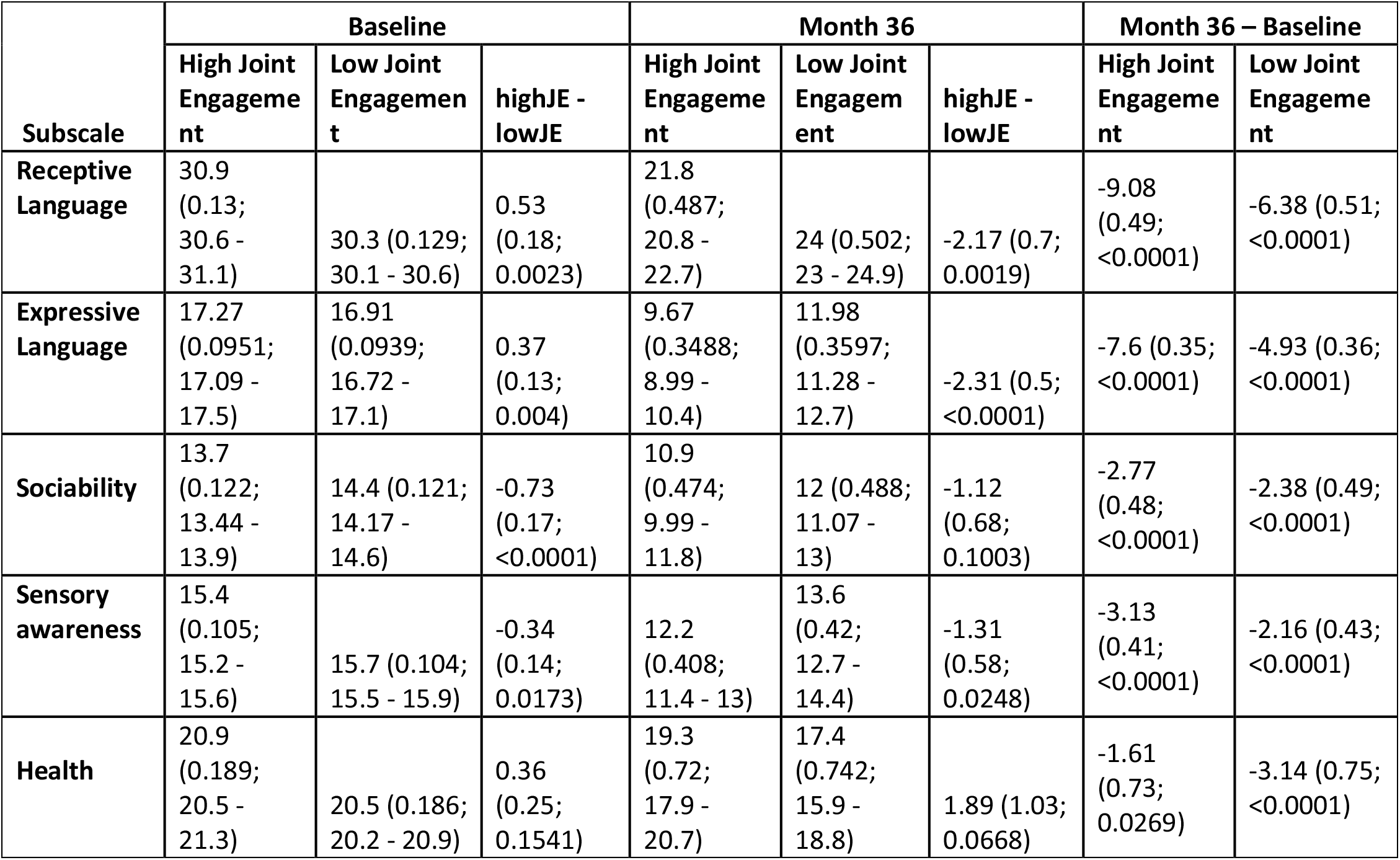
**Characteristics of High Joint Engagement (highJE) and Low Joint Engagement groups (lowJE). Data is presented as LS Means (SE; 95% CI). A lower score indicates a lower severity of ASD symptoms. The difference between the High Joint Engagement and the Low Joint Engagement groups (highJE - lowJE) is presented as: LS Mean (SE; P-value). The negative highJE - lowJE difference indicates that the lowJE group had a higher score and therefore more severe symptoms.**

**Figure 1.**
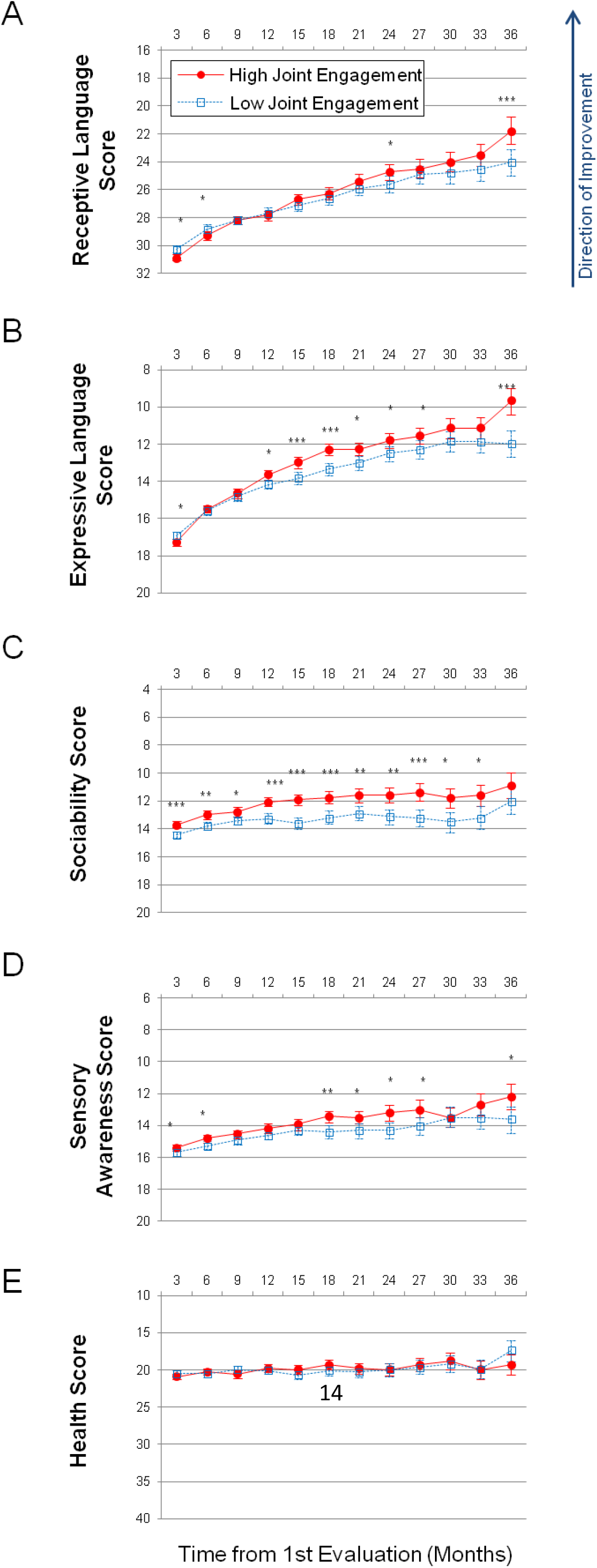
**Longitudinal plots of subscale scores LS Means. Horizontal axis shows months from the 1st evaluation (0 to 36 months). Error bars show the 95% confidence interval. To facilitate comparison between subscales, all vertical axes’ ranges have been normalized to show 40% of their corresponding subscale’s maximum available score. A lower score indicates symptom improvement. P-value is marked: ***<0.0001; **<0.001; *<0.05. (A) Receptive Language score. (B) Expressive Language score. (C) Sociability score. (D) Sensory Awareness score (E) Health score.**

On the Expressive Language subscale, the participants in the highJE group improved over the 36-month period by 7.6 points (SE=0.35, p<0.0001) compared to 4.93 points (SE=0.36, p<0.0001) improvement in the lowJE group, Figure 1B, Table S2. The difference in the highJE group relative to the lowJE group at Month 36 was: -2.31 points (SE=0.5, p<0.0001).

On the Sociability subscale, the subjects in the highJE group improved over the 36-month period by 2.77 points (SE=0.48, p<0.0001) compared to 2.38 points (SE=0.49, p<0.0001) improvement in lowJE group, Figure 1C, Table S3. The difference in the highJE group relative to the lowJE group at Month 36 was insignificant: -1.12 points (SE=0.68, p=0.1003).

On the Sensory Awareness subscale, the subjects in the highJE group improved over the 36-month period by 3.13 points (SE=0.41, p<0.0001) compared to 2.16 points (SE=0.43, p<0.0001) improvement in the lowJE group, Figure 1D, Table S4. The difference in the highJE group relative to the lowJE group at Month 36 was: -1.31 points (SE=0.58, p=0.0248).

On the Health subscale, the subjects in the highJE group improved over the 36-month period by 1.61 points (SE=0.73, p<0.0269) compared to improvement by 3.14 points (SE=0.75, p<0.0001) in the lowJE group, Figure 1E, Table S5. The difference in the highJE group relative to the lowJE group at Month 36 was not statistically significant: 1.89 points (SE=1.03, p<0.0668).

## Discussion

This is the largest study to date demonstrating a strong association between joint engagement and developmental trajectories in children with ASD. Parents assessed the development of 12081 ASD children, two-to six-years-of-age, quarterly for three years. The High Joint Engagement group (highJE) included 2508 children who experienced 3 hours or more of joint engagement per day and the Low Joint Engagement (lowJE) group included 2272 children who experienced 1 hour or less of joint engagement per day. In order to compare the groups, participants in the lowJE group were matched to those in the highJE group using propensity score (Schneider et al., 2007) based on age, gender, expressive language, receptive language, sociability, sensory awareness, and health at the 1^st^ evaluation (baseline). The number of matched participants was 2272 in each group.

Participants in the highJE group improved to a greater extent than matched participants in the lowJE group. On the annualized basis, the highJE group improved their combinatorial receptive language 1.4-times faster (p=0.0019), expressive language 1.5-times faster (p<0.0001), and sensory awareness 1.5-times faster (p=0.0248) than the lowJE group. The difference in sociability and health at the end of 3-year study was insignificant. The results of this study support previous reports of a strong correlation between the joint engagement and language and cognitive development (Fuller et al., 2020).

These findings add significantly to the results of previous studies due to unbiased nature of the collected data. Previous studies investigated effects of proprietary interventions and therefore are in danger of a perceived self-promotion bias. This study does not have any interest in promoting joint engagement as a therapy and, therefore, there cannot have any bias in favor of the intervention. Additionally, the total number of participants in previous studies of joint engagement was in hundreds, while this study reports on the developmental trajectories of 12081 children with ASD.

What were the relationships between joint engagement and other aspects of culture? Very little correlation was detected between the duration of joint engagement and the duration of watching TV and videos (R = 0.05). Furthermore, little correlation was detected between the duration of joint engagement and the duration a child using educational apps on his own (R=0.11). These two observations suggest that joint engagement is not related to TV exposure or the use of electronic devices. Additionally, little correlation was detected between the duration of joint engagement and parents’ education (R=0.06), suggesting that joint engagement is not a function of parental education. Stronger correlation was found with the duration of physical activity (R=0.49) suggesting that parents who spend more time with their children are playing sports with them and pointing to a potential new target for future interventions.

### Limitations

Epidemiological studies of app users provide access to a large number of children at relatively low cost, but have obvious downsides, such as relying on parent reports for diagnosis and assessments. Participants of this study identified their diagnosis as ASD at their last evaluation. Other diagnostic options included: Suspected ASD, Mild Language Delay, Pervasive Developmental Disorder, Attention Deficit Disorder, Social Communication Disorder, Specific Language Impairment, Apraxia, Sensory Processing Disorder, Down Syndrome, Lost Diagnosis of Autism or PDD, Other Genetic Disorder, Normally Developing Child.

Diagnosis misidentification should not have been a significant problem. The app is popular only in the ASD community. Most app users (82%) reported ASD diagnosis by their last evaluation. If ASD participants were mislabeling themselves as a different condition, they would have been excluded from this study. Second, if participants without ASD diagnosis were misidentifying themselves as ASD, they must have also misrepresented their ATEC score (calculated based on 77 questions) as the average initial ATEC total score was 69.0 ± 26.2, which corresponds to moderate-to-severe ASD as reported earlier (Mahapatra, Vyshedsky, et al., 2018) and shown in Table 1. It is highly unlikely that participants went through such an effort to consciously misrepresent their condition since there was no benefit in doing so, although random events of this kind cannot be completely excluded.

We have also previously explored the relationships between different levels of ASD severity reported by parents and the reported scores. We hypothesized that if parents clearly understood and honestly communicated their child’s diagnosis, the reported ASD severity level would have a consistent relationship with assessment subscales. Wherein, greater ASD severity would correspond to worse assessment scores and vice versa. Conversely, if parents misreported their children’s diagnosis, no difference in the average assessment score would be expected between the groups. This cross-sectional analysis of 9573 children has demonstrated statistically significant differences between mild and moderate ASD diagnosis as well as between moderate and severe ASD diagnosis in each subscale and in every age group in children 3 years of age and older (Jagadeesan et al., 2022). These findings contribute to support good fidelity of parents’ reports of children’s diagnosis.

In terms of evaluation scores, two biases are possible. Parents may yield to wishful thinking and overestimate their children’s abilities on a single assessment (Scattone et al., 2011). This, however, is not a problem for a longitudinal study as the pattern of changes generated by measuring the score dynamics over multiple assessments provides meaningful data on a child’s developmental trajectory even if wishful thinking was inflating each individual assessment. A second possible bias concerns parents who may wish to exaggerate their child’s improvement by purposefully entering a score that is better than their previous assessment. If this was the case, one would surmise that parents who have invested more time and energy into the app were also more likely to rate their child as improving. We have previously addressed this possibility analytically by calculating the correlation coefficient between the app usage measured in days/week with child’s improvement in receptive language (*r* = –0.01), expressive language (*r* = –0.06), sociability (*r* = –0.04), cognitive awareness (*r* = –0.01), and health (*r* = 0.01) (Vyshedskiy et al., 2020). Low absolute values of correlation coefficients, as well as the variability of the direction of correlation (positive correlation for improvement of health and negative correlation for other subscales) were not consistent with the hypothesis that parents who invested more time into working with the app were also more likely to rate their child as improving. Additionally, even if parents wanted to score their children as improving (consciously or unconsciously), this would have been very difficult as parents were blinded to their answers at all the previous evaluations and it is near-impossible to recall one’s answers to 133 questions (with each question coming with 3 to 6 options) for three months (time between evaluations). We conclude that it is unlikely that evaluation biases influenced the results of this study.

Another limitation is that this study observational design cannot definitively prove causality since not all confounders can be adjusted appropriately. Our results only inform on the association of joint engagement with better language acquisition and sensory awareness. However, this report in combination with the multitude of randomized controlled studies of joint engagement interventions points to the existence of strong causality between joint engagement and a child’s developmental trajectory.

## Data Availability

All data produced in the present study are available upon reasonable request to the authors.

## Acknowledgements

We wish to thank Dr. Petr Ilyinskii for his scrupulous editing of this manuscript.

## Author contributions

AV and EK designed the study. AV and EK analyzed the data. AV wrote the paper.

## Informed Consent

Caregivers have consented to anonymized data analysis and publication of the results. The study was conducted in compliance with the Declaration of Helsinki (Association, 2013).

## Compliance with Ethical Standards

Using the Department of Health and Human Services regulations found at 45 CFR 46.101(b)(4), the Biomedical Research Alliance of New York LLC Institutional Review Board (IRB) determined that this research project is exempt from IRB oversight.

## Data Availability

De-identified raw data from this manuscript are available from the corresponding author upon reasonable request.

## Code availability statement

Code is available from the corresponding author upon reasonable request.

## Open Practices Statement

The study was preregistered at ClinicalTrials.gov (number NCT02708290).

