## Supplementary Material for "Joint engagement is associated with greater development of language and sensory awareness in children with autism"

**Table S1: LS Means (SE; 95% CI) for Receptive Language MSEC subscale score. The difference between High Joint Engagement and Low Joint Engagement (highJE - lowJE) is presented as: LS Mean (SE; P-value). The negative highJE - lowJE difference indicates that the lowJE group had higher score and therefore more severe symptoms.**

| **Visit Number** | **High Joint Engagement** | **Low Joint Engagement** | **highJE - lowJE** |
| --- | --- | --- | --- |
| Baseline | 30.9 (0.13; 30.6 - 31.1) | 30.3 (0.129; 30.1 - 30.6) | **0.53 (0.18; 0.0023)** |
| Month 6 | 29.3 (0.159; 29 - 29.6) | 28.8 (0.161; 28.5 - 29.1) | **0.53 (0.22; 0.0168)** |
| Month 9 | 28.2 (0.163; 27.9 - 28.5) | 28.2 (0.165; 27.9 - 28.5) | 0.05 (0.23; 0.8291) |
| Month 12 | 27.8 (0.182; 27.5 - 28.2) | 27.7 (0.183; 27.4 - 28.1) | 0.1 (0.25; 0.6902) |
| Month 15 | 26.7 (0.204; 26.3 - 27.1) | 27.1 (0.215; 26.7 - 27.6) | -0.48 (0.29; 0.0988) |
| Month 18 | 26.3 (0.227; 25.8 - 26.7) | 26.6 (0.247; 26.1 - 27.1) | -0.28 (0.33; 0.392) |
| Month 21 | 25.4 (0.261; 24.9 - 25.9) | 25.9 (0.268; 25.4 - 26.5) | -0.57 (0.37; 0.1226) |
| Month 24 | 24.7 (0.292; 24.2 - 25.3) | 25.6 (0.304; 25 - 26.2) | **-0.89 (0.42; 0.0334)** |
| Month 27 | 24.5 (0.322; 23.8 - 25.1) | 24.9 (0.337; 24.2 - 25.5) | -0.38 (0.46; 0.4067) |
| Month 30 | 24 (0.369; 23.3 - 24.7) | 24.8 (0.396; 24 - 25.5) | -0.74 (0.54; 0.1706) |
| Month 33 | 23.5 (0.424; 22.7 - 24.3) | 24.5 (0.426; 23.6 - 25.3) | -0.95 (0.6; 0.1129) |
| Month 36 | 21.8 (0.487; 20.8 - 22.7) | 24 (0.502; 23 - 24.9) | **-2.17 (0.7; 0.0019)** |
| Month 36 - Baseline | -9.08 (0.49; <0.0001) | -6.38 (0.51; <0.0001) | na |

**Table S2: LS Means (SE; 95% CI) for Expressive Language measured by the Subscale 1 of ATEC. The difference between High Joint Engagement and Low Joint Engagement (highJE - lowJE) is presented as: LS Mean (SE; P-value). The negative highJE - lowJE difference indicates that the lowJE group had higher score and therefore more severe symptoms.**

| **Visit Number** | **High Joint Engagement** | **Low Joint Engagement** | **highJE - lowJE** |
| --- | --- | --- | --- |
| Baseline | 17.27 (0.0951; 17.09 - 17.5) | 16.91 (0.0939; 16.72 - 17.1) | 0**.37 (0.13; 0.004)** |
| Month 6 | 15.51 (0.1154; 15.28 - 15.7) | 15.58 (0.1166; 15.35 - 15.8) | -0.07 (0.16; 0.6686) |
| Month 9 | 14.65 (0.1183; 14.42 - 14.9) | 14.82 (0.1198; 14.58 - 15.1) | -0.17 (0.16; 0.3082) |
| Month 12 | 13.65 (0.1316; 13.39 - 13.9) | 14.16 (0.1323; 13.9 - 14.4) | **-0.51 (0.18; 0.0053)** |
| Month 15 | 12.99 (0.1469; 12.71 - 13.3) | 13.85 (0.1551; 13.54 - 14.2) | **-0.85 (0.21; <0.0001)** |
| Month 18 | 12.31 (0.1634; 11.99 - 12.6) | 13.35 (0.1778; 13 - 13.7) | **-1.04 (0.24; <0.0001)** |
| Month 21 | 12.28 (0.1876; 11.91 - 12.6) | 13.02 (0.1925; 12.64 - 13.4) | **-0.74 (0.27; 0.0055)** |
| Month 24 | 11.81 (0.2097; 11.4 - 12.2) | 12.5 (0.2183; 12.07 - 12.9) | **-0.69 (0.3; 0.0219)** |
| Month 27 | 11.56 (0.2308; 11.11 - 12) | 12.3 (0.242; 11.82 - 12.8) | **-0.74 (0.33; 0.0266)** |
| Month 30 | 11.14 (0.2643; 10.62 - 11.7) | 11.86 (0.284; 11.3 - 12.4) | -0.72 (0.39; 0.0633) |
| Month 33 | 11.14 (0.3034; 10.54 - 11.7) | 11.87 (0.3053; 11.28 - 12.5) | -0.74 (0.43; 0.0861) |
| Month 36 | 9.67 (0.3488; 8.99 - 10.4) | 11.98 (0.3597; 11.28 - 12.7) | **-2.31 (0.5; <0.0001)** |
| Month 36 - Baseline | -7.6 (0.35; <0.0001) | -4.93 (0.36; <0.0001) | na |

**Table S3: LS Means (SE; 95% CI) for Sociability subscale score measured by the Subscale 2 of ATEC. The difference between High Joint Engagement and Low Joint Engagement (highJE - lowJE) is presented as: LS Mean (SE; P-value). The negative highJE - lowJE difference indicates that the lowJE group had higher score and therefore more severe symptoms.**

| **Visit Number** | **High Joint Engagement** | **Low Joint Engagement** | **highJE - lowJE** |
| --- | --- | --- | --- |
| Baseline | 13.7 (0.122; 13.44 - 13.9) | 14.4 (0.121; 14.17 - 14.6) | **-0.73 (0.17; <0.0001)** |
| Month 6 | 13 (0.151; 12.71 - 13.3) | 13.8 (0.153; 13.5 - 14.1) | **-0.8 (0.21; 0.0001)** |
| Month 9 | 12.8 (0.155; 12.46 - 13.1) | 13.4 (0.157; 13.09 - 13.7) | **-0.63 (0.22; 0.0035)** |
| Month 12 | 12.1 (0.174; 11.74 - 12.4) | 13.3 (0.175; 12.96 - 13.7) | **-1.23 (0.24; <0.0001)** |
| Month 15 | 11.9 (0.195; 11.55 - 12.3) | 13.6 (0.207; 13.18 - 14) | **-1.66 (0.28; <0.0001)** |
| Month 18 | 11.8 (0.218; 11.32 - 12.2) | 13.2 (0.238; 12.78 - 13.7) | **-1.49 (0.32; <0.0001)** |
| Month 21 | 11.6 (0.252; 11.09 - 12.1) | 12.9 (0.258; 12.41 - 13.4) | **-1.33 (0.36; 0.0002)** |
| Month 24 | 11.6 (0.282; 11.03 - 12.1) | 13.1 (0.294; 12.49 - 13.6) | **-1.48 (0.4; 0.0003)** |
| Month 27 | 11.4 (0.312; 10.74 - 12) | 13.2 (0.327; 12.55 - 13.8) | **-1.84 (0.45; <0.0001)** |
| Month 30 | 11.8 (0.358; 11.13 - 12.5) | 13.5 (0.385; 12.71 - 14.2) | **-1.63 (0.52; 0.0018)** |
| Month 33 | 11.6 (0.412; 10.83 - 12.4) | 13.2 (0.414; 12.4 - 14) | **-1.57 (0.58; 0.0068)** |
| Month 36 | 10.9 (0.474; 9.99 - 11.8) | 12 (0.488; 11.07 - 13) | -1.12 (0.68; 0.1003) |
| Month 36 - Baseline | -2.77 (0.48; <0.0001) | -2.38 (0.49; <0.0001) | na |

**Table S4: LS Means (SE; 95% CI) for the Sensory/Cognitive Awareness subscale score measured by the Subscale 3 of ATEC. The difference between High Joint Engagement and Low Joint Engagement (highJE - lowJE) is presented as: LS Mean (SE; P-value). The negative highJE - lowJE difference indicates that the lowJE group had higher score and therefore more severe symptoms.**

| **Visit Number** | **High Joint Engagement** | **Low Joint Engagement** | **highJE - lowJE** |
| --- | --- | --- | --- |
| Baseline | 15.4 (0.105; 15.2 - 15.6) | 15.7 (0.104; 15.5 - 15.9) | **-0.34 (0.14; 0.0173)** |
| Month 6 | 14.8 (0.13; 14.6 - 15.1) | 15.3 (0.132; 15.1 - 15.6) | **-0.47 (0.18; 0.0093)** |
| Month 9 | 14.5 (0.134; 14.3 - 14.8) | 14.9 (0.135; 14.6 - 15.1) | -0.33 (0.19; 0.0797) |
| Month 12 | 14.2 (0.15; 13.9 - 14.5) | 14.6 (0.15; 14.3 - 14.9) | -0.41 (0.21; 0.0508) |
| Month 15 | 13.9 (0.168; 13.6 - 14.3) | 14.3 (0.178; 14 - 14.7) | -0.42 (0.24; 0.0831) |
| Month 18 | 13.4 (0.188; 13.1 - 13.8) | 14.4 (0.205; 14 - 14.8) | **-1 (0.27; 0.0003)** |
| Month 21 | 13.5 (0.217; 13.1 - 13.9) | 14.3 (0.222; 13.8 - 14.7) | **-0.77 (0.31; 0.0123)** |
| Month 24 | 13.2 (0.243; 12.7 - 13.7) | 14.3 (0.253; 13.8 - 14.8) | **-1.06 (0.35; 0.0024)** |
| Month 27 | 13 (0.268; 12.4 - 13.5) | 14 (0.281; 13.4 - 14.5) | **-1.01 (0.39; 0.0087)** |
| Month 30 | 13.5 (0.308; 12.9 - 14.1) | 13.5 (0.331; 12.9 - 14.2) | 0.04 (0.45; 0.9377) |
| Month 33 | 12.7 (0.354; 12 - 13.4) | 13.5 (0.356; 12.8 - 14.2) | -0.74 (0.5; 0.138) |
| Month 36 | 12.2 (0.408; 11.4 - 13) | 13.6 (0.42; 12.7 - 14.4) | **-1.31 (0.58; 0.0248)** |
| Month 36 - Baseline | -3.13 (0.41; <0.0001) | -2.16 (0.43; <0.0001) | na |

**Table S5: LS Means (SE; 95% CI) for Health/Physical/Behavior subscale score measured by the Subscale 4 of ATEC. The difference between High Joint Engagement and Low Joint Engagement (highJE - lowJE) is presented as: LS Mean (SE; P-value). The negative highJE - lowJE difference indicates that the lowJE group had higher score and therefore more severe symptoms.**

| **Visit Number** | **High Joint Engagement** | **Low Joint Engagement** | **highJE - lowJE** |
| --- | --- | --- | --- |
| Baseline | 20.9 (0.189; 20.5 - 21.3) | 20.5 (0.186; 20.2 - 20.9) | 0.36 (0.25; 0.1541) |
| Month 6 | 20.3 (0.232; 19.8 - 20.7) | 20.5 (0.235; 20 - 21) | -0.25 (0.32; 0.4431) |
| Month 9 | 20.6 (0.238; 20.1 - 21.1) | 20 (0.241; 19.6 - 20.5) | 0.56 (0.33; 0.0925) |
| Month 12 | 19.8 (0.266; 19.2 - 20.3) | 20.1 (0.268; 19.6 - 20.6) | -0.32 (0.37; 0.3882) |
| Month 15 | 20 (0.298; 19.4 - 20.6) | 20.7 (0.316; 20.1 - 21.3) | -0.69 (0.43; 0.1057) |
| Month 18 | 19.3 (0.333; 18.6 - 19.9) | 20.1 (0.363; 19.4 - 20.9) | -0.84 (0.49; 0.0826) |
| Month 21 | 19.8 (0.384; 19.1 - 20.6) | 20.3 (0.394; 19.6 - 21.1) | -0.5 (0.54; 0.3539) |
| Month 24 | 20 (0.43; 19.1 - 20.8) | 20 (0.448; 19.1 - 20.9) | -0.03 (0.62; 0.9649) |
| Month 27 | 19.3 (0.474; 18.4 - 20.2) | 19.7 (0.497; 18.8 - 20.7) | -0.44 (0.68; 0.5213) |
| Month 30 | 18.8 (0.544; 17.7 - 19.9) | 19.2 (0.585; 18.1 - 20.3) | -0.4 (0.79; 0.6182) |
| Month 33 | 20 (0.626; 18.8 - 21.2) | 19.9 (0.629; 18.7 - 21.1) | 0.1 (0.88; 0.9139) |
| Month 36 | 19.3 (0.72; 17.9 - 20.7) | 17.4 (0.742; 15.9 - 18.8) | 1.89 (1.03; 0.0668) |
| Month 36 - Baseline | -1.61 (0.73; 0.0269) | -3.14 (0.75; <0.0001) | na |
